# G6PD Deficiency Overrepresented Among Pediatric COVID-19 Cases in One Saudi Children Hospital

**DOI:** 10.1101/2020.07.08.20148700

**Authors:** Maryam Al-Aamri, Fatima Al-Khalifa, Fawatim Al-Nahwi, Heba Al-Ameer, Sameer Al-Abdi

## Abstract

Fluorescent spot test for glucose-6-phosphate dehydrogenase (G6PD) deficiency was performed in 5 boys and 14 girls who had confirmed COVID-19. Out of those, 4 (80%) boys and 5 (36%) girls were found to be G6PD deficient.

## Main Text

On March 12, 2020, the first case of coronavirus disease 19 (COVID-19) was diagnosed in the Al-Ahsa area, Saudi Arabia [1]. Until the end of April 2020, all children of less than 14 years old with confirmed COVID-19 were admitted to the Maternity and Children hospital in Al-Ahsa area. During this period, 29 children (8 boys and 21girls) were admitted. All these cases were either asymptomatic or with a mild COVID-19. Fluorescent spot test (FST) for glucose-6-phosphate dehydrogenase (G6PD) deficiency was performed in 5 boys and 14 girls. Out of those, 4 (80%) boys and 5 (36%) girls were found to be G6PD deficient. Our FST has a cut-off point of 2.1 Units/gram Hemoglobin, which indicates these cases are moderate to severe G6PD deficiency. All these cases were either asymptomatic or with a mild COVID-19. The G6PD deficiency is overrepresented in this small case series as the prevalence of G6PD deficiency in Al-Ahsa is 23% in males and 13% in females [2]. We and others anticipate that G6PD deficient individuals will be more vulnerable to severe acute respiratory syndrome coronavirus 2, the causative agent of the COVID-19 pandemic [3, 4]. Still, this needs to be confirmed in a large-scale population-based study.

## Data Availability

Data are available upon request.

